# Impact of Genetic Background as a Risk Factor for Atherosclerotic Cardiovascular Disease: A Protocol for a Nationwide Genetic Case-Control (CV-GENES) study in Brazil

**DOI:** 10.1101/2023.07.19.23292905

**Authors:** Antônio José Cordeiro Mattos, Precil Diego Miranda de Menezes Neves, Gustavo Bernardes de Figueiredo Oliveira, Frederico Rafael Moreira, Maria Carolina Pintão, Viviane Zorzanelli Rocha, Cristiane de Souza Rocha, Viviane Nakano Katz, Elisa Napolitano Ferreira, Diana Rojas-Málaga, Celso Ferraz Viana, Fabiula Fagundes da Silva, Juliete Jorge Vidotti, Natalia Mariana Felicio, Leticia de Araujo Vitor, Karina Gimenez Cesar, Camila Araújo da Silva, Lucas Bassolli de Oliveira Alves, Haliton Alves de Oliveira Junior, Álvaro Avezum

## Abstract

Atherosclerotic Cardiovascular Disease (ASCVD) represents the leading cause of death worldwide, and individual screening should be based on behavioral, metabolic, and genetic profile derived from data collected in large population-based studies. Due to a polygenic nature of ASCVD, we aimed to assess the association of genomics to ASCVD risk and its impact on the occurrence of acute myocardial infarction, stroke, or peripheral artery thrombotic-ischemic events on a population level. CV-GENES is a multicenter, Brazilian nationwide, 1:1 case-control study of 3,734 patients. Inclusion criteria for cases are the first occurrence of one of the cardiovascular events. Individuals without known ASCVD, and age- and sex-matched will be eligible for the control group. A genetics core lab analysis will be performed through the association of low-pass whole genome sequencing and whole exome sequencing. A polygenic risk score will be built in a multiethnic population to estimate the association between genetic polymorphisms and risk of ASCVD. In addition, the presence of pathogenic or likely pathogenic variants will be screened in 8 genes (*ABCG5*, *ABCG8*, *APOB*, *APOE, LDLR*, *LDLRAP1*, *LIPA, PCSK9*) associated with atherosclerosis. Multiple logistic regression will be applied to estimate adjusted odds ratios (ORs) and 95% confidence intervals (CIs), and population attributable risks will be calculated. This study is registered in clinicaltrials.gov (NCT05515653.)

## Introduction

Atherosclerotic cardiovascular disease (ASCVD) represents the leading cause of death worldwide. One third of overall deaths is associated to cardiovascular disease.[1,2] Documenting the consistency or variations in the associations between risk factors and both cardiovascular disease (CVD) and mortality, both globally and in countries grouped by income levels, will help the development of global and context-specific strategies for prevention. International cohort findings indicate that over 70% of cardiovascular disease cases can be attributed to a small cluster of modifiable risk factors. [3]

Low- and middle-income countries are substantially affected by CVD. In Brazil, CVD are responsible for more than 300,000 deaths/year, representing the main cause of death, followed by cancer, respiratory diseases, and diabetes. Together, chronic non-communicable diseases (NCDs) are responsible for approximately 70% of the causes of death in both sexes. [1,2,4,5]

Contemporary studies have shown that seven out of ten cases of CVD can be explained by easy to identify and modifiable risk factors such as hypertension, low level of education, smoking, dyslipidemia, unhealthy diet, abdominal obesity, sedentary lifestyle, diabetes, psychosocial factors, and air pollution.[3] However, recently, advances in genetic sequencing technologies associated with a greater access to them led to the identification of genetic polymorphisms associated with an increased risk of CVD, emphasizing the model of interaction between genetics and environment as a trigger for CVD but, unlike other known risk factors, the genetic background is a non-modifiable factor.[6,7]

The Brazilian population is unique due to its ethnic diversity, which may impact on the risk of specific diseases, including CVD. [8,9] The impact of the genetic component on the susceptibility to illnesses in the Brazilian population has already been shown for infectious, autoimmune, and hematological diseases, in addition to drug pharmacokinetics and even on the transplant allocation system. [7,10-15]

From a public health system perspective and epidemiological characteristics on cardiovascular health in developing countries, several factors have been already described.[16,17], such as the need for robust studies with national representation which allow the knowledge of the epidemiological peculiarities and genetic contributors to CVD. To reliably answer this question, a genetic case-control study (CV-GENES) was designed to evaluate the contribution of genetic background adjusted for traditional modifiable cardiovascular risk factors, in a nationwide Brazilian cohort.[18,19]

The primary aim of the current study is to assess the association of genomics to ASCVD risk and its impact on the occurrence of acute myocardial infarction, stroke, or peripheral artery thrombotic-ischemic events on a population level.

## Materials and Methods

### Protocol report

CV-GENES protocol version 2.0/2022 was reported through an adaptation of the *Standard Protocol Items: Recommendations for Interventional Trials* (SPIRIT). **Fig. 1** shows the study schedule of enrolment, and assessments. The SPIRIT checklist is reported in **Supplementary Material 1**. Protocol in original language and its English-translated version is in **Supplementary Material 2**.

**Fig 1.**
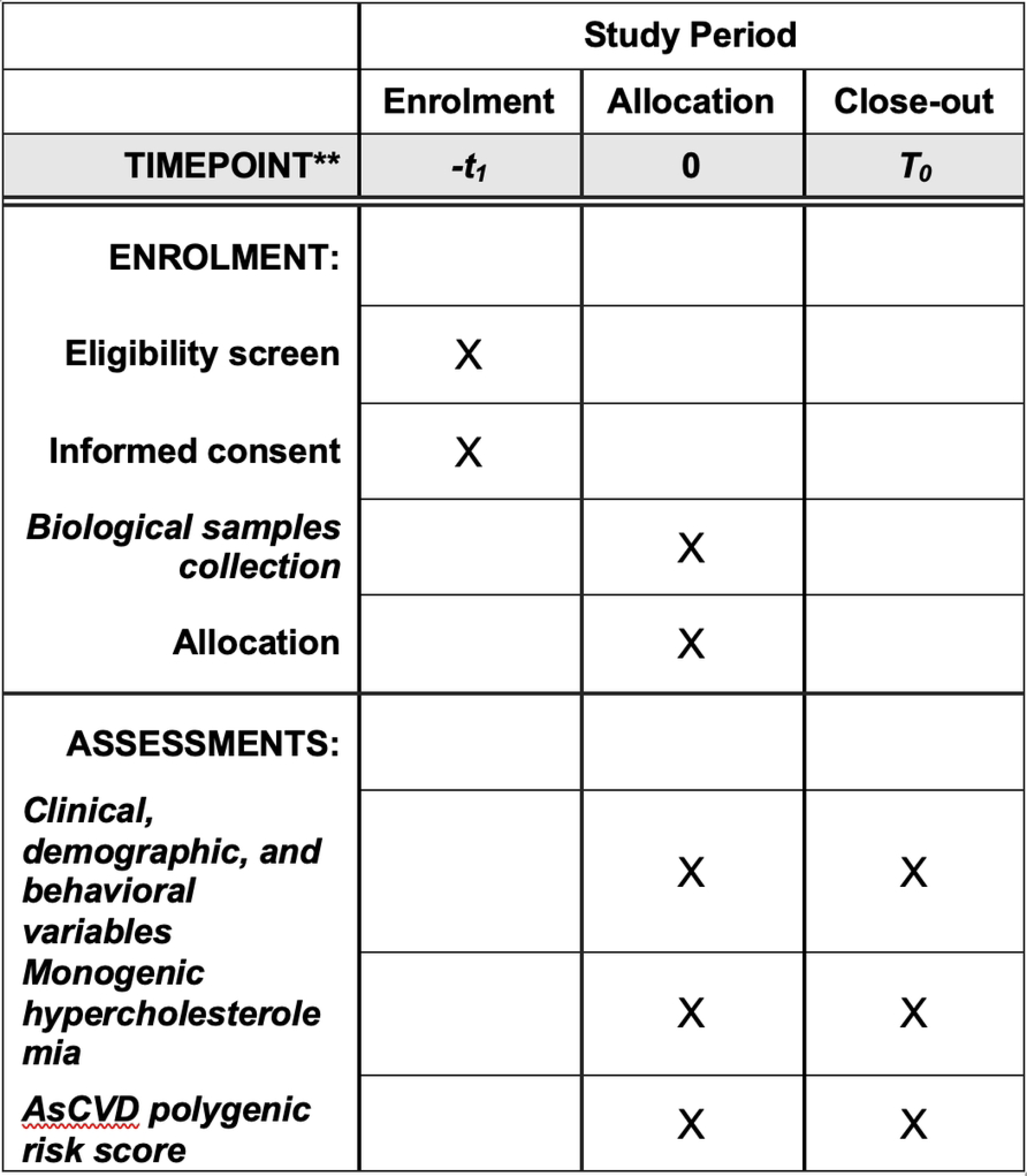
SPIRIT schedule for study enrollment and assessments.

### Study Design

A multicenter observational case-control study will be conducted at 50 centers from all geographical regions across Brazil to account for ethnical, cultural, and socioeconomic aspects.

### Patient eligibility

All patients ≥18 years of age admitted to hospital due to the first occurrence of an acute myocardial infarction (AMI), stroke, or peripheral artery thrombotic event, presenting between 1-5 days of symptom onset, will be screened (**Supplementary Material 3**).

Eligible cases must have signs and/or symptoms compatible with the acute manifestation of ASCVD in one of the arterial territories. Case definitions include clinical and diagnostic criteria and are based on national and international guidelines. Acute myocardial infarction is defined as: clinical presentation, electrocardiogram patterns (new pathological Q waves, and/or elevation of ST segment of 1 mm in ≥ 2 contiguous leads, or 2 mm from V1 to V3, or new left bundle branch block, or new changes of the ST-T segment, and cTnI or cTnT curve compatible with acute myocardial injury of ischemic nature (which is currently named a type 1 acute MI); stroke is defined as clinical signs of rapid development of focal or global brain function disorder, lasting more than 24 hours, with no apparent cause other than that of vascular origin (does not include trauma, cancer, infection of the central nervous system), CT or MRI with positive results; and peripheral arterial thrombotic event, as clinical presentation (pain, burning sensation, absence of pulse, decreased local temperature, changes in color) and diagnostic tests (Doppler ultrasound, angiotomography, or arteriography) that show interruption of flow to the lower limbs acutely, of embolic or thrombotic cause, leading to distal ischemia.[20-23]

Controls will comprise patients seeking medical care for non-ASCVD conditions, thus will be enrolled during hospitalization due to other causes, or also healthy individuals with no clinical complain, from the same community. Cases and controls will be recruited in a 1:1 ratio, adjusted for sex and age.

Exclusion criteria for both cases and controls are previously known ASCVD, e.g., chronic coronary artery disease, stable angina, previous myocardial infarction, percutaneous revascularization procedures (coronary, cerebrovascular, or peripheral), revascularization surgery in any arterial vascular bed, transient ischemic attack, stroke, aortic aneurysm, intermittent claudication, peripheral artery disease that led to amputation.

### Recruitment

Strategies to reinforce the engagement of research teams in all centers towards optimizing recruitment include: payment of a fee for each inclusion; return of biochemical and genetic tests to patients; taking advantage of seasonality (colder seasons have a higher incidence of AsCVD); close monitoring of recruitment rates; weekly meetings with participating centers.

### Sample size calculation

For the sample size calculation, we applied the following assumptions: odds ratio for genetics was set at 1.3, allele frequency of 10%, inheritance model was dominant model (since the presence of only one polymorphic allele is necessary to the risk calculation), the prevalence of cardiovascular disease in the general population of 10%, statistical power of 90%, type I error rate (α) of 0.05, 2-sided significance tests, resulting in a sample of 1,867 cases and 1,867 controls. Sample-size calculation was performed using Quanto Software Version 1.2.4.

### Variables

The exposure to modifiable risk factors in combination with genomic data (polygenic risk score) in both cases and controls will be expressed as odds ratios (95% CI). Multiple logistic regression models will be performed to adjust and determine the strength of association between demographic variables (sex, age, race), traditional risk factors (smoking, diabetes, hypertension, obesity, anxiety and depression, unhealthy diet, physical inactivity, alcohol consumption, and apolipoprotein B/A1 ratio (ApoB/ApoA1)), and genetic data (Polygenic Risk Score, PRS). For each variable with a higher chance of cardiovascular disease (significant OR), attributable risk will be calculated for the genetic component (PRS) and to other covariables. **Fig 2** shows the Polygenic Risk Score flowchart.

**Fig 2.**
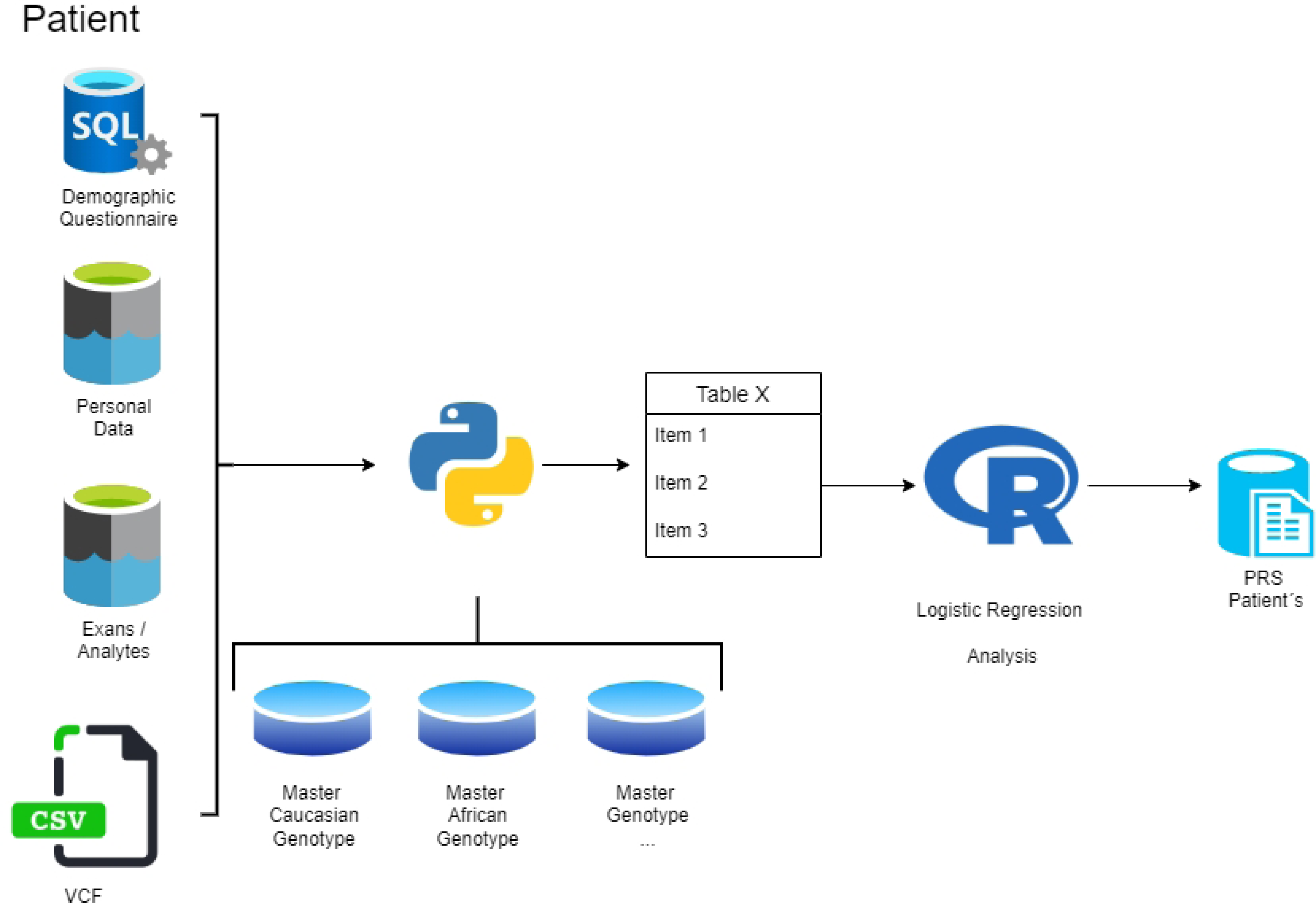
Polygenic Risk Score flowchart.

**Table 1** exhibits the biochemical tests and its respective stability and methodology. All the biochemical and genetic tests will be run in the same centralized core lab.

**Table 1:** Biochemical tests and its respective stability and methodology.

| Test description | Refrigerated Stability | Frozen Stability | Methodology |
| --- | --- | --- | --- |
| Glycosylated hemoglobin, whole blood | (2-8 °C): 7 days; | (-20 °C): 30 days; | Ion exchange Column chromatography in HPLC system |
| Apolipoprotein A-1, serum | (2-8 °C): 8 days; | (-20 °C): 2 months; | Immunoturbidimetric |
| Apolipoprotein B, serum | (2-8 °C): 8 days; | (-20 °C): 2 months; | Immunoturbidimetric |
| Cholesterol, serum | (2-8 °C): 7 days; | (-20 °C): 3 months; | Colorimetric enzyme |
| HDL Cholesterol, serum | (2-8 °C): 7 days; | (-20 °C): 3 months; | Colorimetric enzyme |
| Cholesterol, LDL Fraction, serum | (2-8 °C): 7 days; | (-20 °C): 3 months; | Calculation based on Friedewald formula |
| Cholesterol, VLDL Fraction, serum | (2-8 °C): 7 days; | (-20 °C): 3 months; | Martin and colleagues' formula |
| Triglycerides, serum | (2-8 °C): 7 days; | (-20 °C): 1 year; | Colorimetric enzyme |
| Creatinine, serum | (2-8 °C): 5 days; | (-20 °C): 1 year. | Colorimetric kinetic |
| Sodium, serum | (2-8 °C): 7 days; | (-20 °C): 6 months; | Potentiometric |
| Potassium, serum | (2-8 °C): 7 days; | (-20 °C): 6 months; | Potentiometric |
| Sodium, spot, urine | (2-8 °C): 7 days; | (-20 °C): 6 months; | Potentiometric |
| Potassium, urine | (2-8 °C): 7 days; | (-20 °C): 3 months; | Potentiometric |
| Creatinine, spot, urine | (2-8 °C): 5 days; | (-20 °C): 1 year; | Colorimetric kinetic |
| Albuminuria, spot, urine | (2-8 °C): 14 days; | (-20 °C): 6 months; | Immunoturbidimetric assay |

### Data source and measurement

Data will be collected through electronic case report forms (eCRFs) specifically designed for this study by using the REDCap software.[24] Demographic variables, prior and inpatient use of medications, laboratory tests (glycated hemoglobin, serum apolipoprotein A-1, serum apolipoprotein B, serum cholesterol, total serum cholesterol and fractions [HDL-c, LDL-c, and VLDL-c], serum triglycerides, serum sodium, serum potassium, and measurement of sodium, potassium, creatinine, and albumin in a spot urine sample), and the genetics results (PRS) will be collected.

The genetic evaluation will be performed through the association of low-pass whole genome sequencing (WGS) (coverage 0.5-1x) with data imputation and whole exome sequencing (WES) (average coverage 90x). The method of PRS calculation is described in the Supplementary Methods. In addition, the presence of pathogenic or likely pathogenic variants will be screened in 8 genes (*ABCG5*, *ABCG8*, *APOB*, *APOE, LDLR*, *LDLRAP1*, *LIPA, PCSK9*) associated with atherosclerosis, from the WES (**Supplementary material 3**).

## Data analysis

### Quantitative variables and statistical analysis

The Hardy-Weinberg genetic equilibrium test will be evaluated in the control group using the chi-square test or Fisher’s exact test. To estimate the association between genetic polymorphisms and risk of cardiovascular disease, univariate and multiple unconditional logistic regression analyses will be conducted. [25-29]

Initially, univariate binary logistic regression analyses will be performed. Thus, covariates with a p-value < 0.20 in univariate regression analyses will be considered in multiple logistic regression analysis with selection of variables according to the backward elimination technique.[28,29] In backward selection, the selection process in the Backward elimination allows the sample size to adjust and always uses the maximum available sample as the number of variables in the model drops.[30] Occasionally, covariates judged as confounding factors by the investigator may be forced into the final backward regression model.

Additional multiple logistic regression analyses may also be conducted using a variant of the Purposeful Selection algorithm described by Bursac *et al.*[31]

The assumption of linearity on the logit scale (log-odds) between each quantitative covariate and the binary response variable in binary logistic regression analysis will be evaluated with the construction of “Smoothed Scatter Plots” and the method of fractional polynomials. [28,32] When the assumption is not satisfied, quantitative covariates will be categorized for use in logistic regression using cut-off points, according to the literature, distribution tertiles or optimal cut-off obtained from the Receiver Operating Characteristic (ROC) curve, whichever is deemed most appropriate. In case of an optimal cut-off point, it will be defined as the one that maximizes the Youden index.[32,33]

### Estimation of Population Attributable Risk

To characterize the study population, categorical variables will be described as counts and proportions and compared with Pearson’s chi-square test or with Fisher’s exact test. [34] Normally and asymmetrically distributed quantitative variables will be expressed as mean (standard deviation) or median (interquartile range), respectively. [35] Normality will be assessed by visually inspecting histograms and applying normality tests, if appropriate. [36,37] Comparison of these continuous type variables will be carried out with Student’s t test for independent samples or Mann-Whitney test for non-parametric distribution. [34]

All statistical analyses will follow the complete case analysis principle. All hypothesis tests will be 2-sided and p-value<0.05 considered statistically significant. Statistical data analysis will be conducted with SAS 9.4 (SAS Institute, Cary, NC).

### Bias control

Eligibility criteria for cases and controls will be strictly followed to reduce selection bias. The criteria defined for the first ASCVD event was standardized, and all sites will be trained for the correct identification, data recording, and reporting of information. Cases will be validated by the adjudication committee for an index event through clinical data, complementary exams, hospitalization summary, among others, using the methodology applied and validated for diagnoses [38,39]. The relationship between the variables will be adjusted by multivariate logistic models. Data monitoring will be applied throughout the study to assure data quality. Blood and urine samples collect will be standardized, and genetic and biochemistry analysis will be performed in a core lab to guarantee the standardization of methods and quality control.

### Study organization

The Hospital Alemão Oswaldo Cruz is responsible for the national coordination, clinical monitoring and data quality assurance, and adjudication of cases and controls. All biochemistry and genetics analyzes will be conducted by Fleury Group S.A.

### Ethics Statement

This study has been approved by the Institutional Review Board (IRB) at Hospital Alemão Oswaldo Cruz and by all local Ethics Committees at each participating site (CAAE - 56482922.2.1001.0070), on April 22^nd^, 2022. Any changes will be treated as protocol amendments and will be immediately submitted to IRB. IRB approval letter is shown in **Supplementary material 4.**

All patients need to sign two informed consent forms (general study and biobank), which will be applied to both cases and control in each research center by delegated researchers and physicians. A specific anonymized id will be assigned to each participant and their biological samples. Data will be stored in a secure and encrypted cloud, and all patient data will be totally anonymized.

This study is registered in clinicaltrials.gov (NCT05515653) and is currently recruiting. First patient was recruited in July 2022. The estimated completion date is April 2024.

## Discussion

We have designed and launched a nationwide multicenter observational case-control study to evaluate the genetic contribution to ASCVD. We will be able to evaluate the polygenic contribution to ASCVD and, furthermore, to evaluate a set of specific monogenic inheritance genes associated with atherosclerosis in the most ethnic-diverse population already known. This national effort will provide genetic data to support the National Program in Genomics and Precision Medicine (GENOMAS BR), which has the aim to screen 100.000 Brazilians.

Cardiovascular events worldwide are most associated to nine modifiable risk factors (ApoB/ApoA1 ratio, hypertension, diabetes, obesity, smoking, psychosocial factors, poor diet, regular alcohol consumption and regular physical activity), which account for 90% of the population attributable risk (PAR) of both acute myocardial infarction and stroke. Therefore, it is biologically plausible that the genetic background might also play a role in the remaining 10% PAR of ASCVD.

In this study, the contribution of the genetic background in the risk of ASCVD in Brazilian individuals will be ascertained by the analysis of variants with a potential direct effect of disease manifestation (monogenic diseases). The polymorphisms will be ascertained in genes that compound the risk in association to each other (polygenic diseases) that behave as factors of susceptibility to a disease, in the “two hits” pathogenic model, where there is a direct interaction of a genetic background with environmental factors for the development of the disease. [8-10]

A previous study of 8,795 individuals of European, South Asian, Arab, Iranian, and Nepalese origin from the INTERHEART case-control study performed a genotyping of 1,536 single-nucleotide polymorphisms (SNPs) from 103 genes evaluated as well as analyzed previous established cardiovascular risk factors (APOB/A1 ratio, hypertension, diabetes, abdominal obesity, smoking, sedentary lifestyle, alcoholism, and depression). Thirteen polymorphisms were associated with increased cardiovascular risk, 11 of which were related to serum levels of APOB/A1 ratio, 1 associated to LDL-cholesterol receptor and 1 to Apolipoprotein E.[7]

The INTERHEART study showed that genetic risk markers for the occurrence of acute myocardial infarction (AMI), identified through genome analysis, appear to be broadly associated with AMI in several different ethnic groups. However, the population attributable risk to these genetic factors is relatively small compared to modifiable risk factors. In the Latin American subgroup, the impact of the genetic risk score was PAR 0.86 95%CI 0.74-0.93 and PAR 0.12 95%CI 0.02-0.42, respectively, for modifiable and non-modifiable factors. However, as already mentioned, there are no data exclusively from Brazilian individuals with adequate statistical power and standardized data collection of traditional risk factors and, therefore, recommended for an integrated assessment of the PRS impact on the population attributable risk for the occurrence of atherosclerotic events in its broad clinical spectrum.

In a review of the main studies of susceptibility loci for coronary artery disease, including those from important consortia such as CARDIoGRAM, MIGen, WTCCC and Cardiogenics, deCODE, CARDIoGRAM, C4D and CARDIoGRAM + C4D,[40] important contributions were derived from the identification of 60 loci, whose mechanism of action is related to serum levels of LDL-cholesterol, lipoprotein (a) and triglycerides, blood pressure, obesity, coagulation profile, changes in endothelial and smooth muscle cells of the vascular wall, migration mechanisms and cell adhesion, immune activation, inflammation, cell growth, differentiation and apoptosis, as well as extracellular matrix constituents and also some loci with unknown function.

Polymorphisms in genes associated with the regulation of calcium levels (CASR, CYP24A1, CARS, DGKD, DGKH/KIAA0564 and GATA3) [41] and metalloproteinases (MMP-3 and MMP-9) [42] were also associated with an increased risk of coronary artery disease and AMI. The list of polymorphisms associated with AMI progressively increases, and some authors have already reinforced the importance of using the genetic background in cardiovascular risk calculators.[43-45]

Regarding stroke cases, polymorphisms in some genes such as MTHFR, eNOS, ACE, AGT, ApoE, PON1, PDE4D were associated with a higher risk of ischemic stroke. For cases of hemorrhagic stroke, polymorphisms in collagen genes, TLR4 and CD14 and even the gene that gives rise to C-reactive protein were identified. [46]

Studies have shown that genetic background may exert a significant effect on increased risk of cardiovascular disease.[47] In a mixed population such as ours, determining the attributable risk may lead to the development of actions aimed at specific subpopulations, such as providing appropriate genetic advice and prevention strategies. This is particularly important considering that lifestyle changes and even correct adherence to pharmacological therapies are not implemented or followed in a sustained way.

The contribution of the genetic component on the pattern or susceptibility to diseases in the Brazilian population has already been shown to have an impact on the control and treatment of infectious, autoimmune, and hematological diseases, drug pharmacokinetics, and even on the transplant allocation system. [13,14,19,48,49]

Knowing the true impact on the Brazilian population may prompt the implementation of population genetic screening programs for strict control of cardiovascular risk factors and/or indicate changes in therapeutic targets and clinical control.

In this context, the detection of polymorphisms associated with cardiovascular disease could identify patients where modifiable risk factors can be screened/treated early, aiming at cardiovascular prevention. In the same way, the results might help design and implement specific screening aimed at early detection and prompt risk factors control to reduce the burden of premature cardiovascular morbidity and mortality among those with higher PRS combined with cluster of modifiable risk factors.

## Data Availability

No datasets were generated or analysed during the current study. All relevant data from this study will be made available upon study completion

## Acknowledgments

This study was conceived with the support of the Brazilian Unified Health System Institutional Development Program (PROADI-SUS).

## Author contributions

Conception and design of the research: AJCM, PDMMN, GBFO, Oliveira HAOJ, AA. Acquisition of data: MCP, VZR, CSR, VNK, ENF, DR-M, CFV, FFS, JJV, NMF, LAV, KGC, CAS. Statistical analysis plan: FRM, LBOA. Obtaining financing: GBFO, CAS, HAOJ, AA. Writing of the manuscript: AJCM, PDMMN, GFO, MCP, VNK FRM, HOJ, AA. Critical revision of the manuscript for important intellectual content: AJCM, PDMMN, GBFO, FRM, MCP, VZR, CSR, VNK, HAOJ, AA.

## Data Sharing

Anonymized data, study protocol, statistical analysis plan, and informed consent form will be shared upon reasonable request to the corresponding author. All patients and research centers will be communicated about the study results. Patients will be informed about the genetic tests result and genetic counselling might be implemented.

## REFERENCES

1. Collaborators GCoD (2018) Global, regional, and national age-sex-specific mortality for 282 causes of death in 195 countries and territories, 1980-2017: a systematic analysis for the Global Burden of Disease Study 2017. Lancet 392: 1736-1788.

2. Collaborators GRF (2018) Global, regional, and national comparative risk assessment of 84 behavioural, environmental and occupational, and metabolic risks or clusters of risks for 195 countries and territories, 1990–2017: a systematic analysis for the Global Burden of Disease Study 2017. Lancet (London, England) 392: 1923.

3. Yusuf S, Joseph P, Rangarajan S, Islam S, Mente A, et al. (2020) Modifiable risk factors, cardiovascular disease, and mortality in 155 722 individuals from 21 high-income, middle-income, and low-income countries (PURE): a prospective cohort study. The Lancet 395: 795–808.

4. O’Donnell MJ, Chin SL, Rangarajan S, Xavier D, Liu L, et al. (2016) Global and regional effects of potentially modifiable risk factors associated with acute stroke in 32 countries (INTERSTROKE): a case-control study. Lancet 388: 761–775.

5. Yusuf S, Hawken S, Ounpuu S, Dans T, Avezum A, et al. (2004) Effect of potentially modifiable risk factors associated with myocardial infarction in 52 countries (the INTERHEART study): case-control study. Lancet 364: 937–952.

6. Genômica SBdGMe (2020) PARECER TÉCNICO SOCIEDADE BRASILEIRA DE GENÉTICA MÉDICA E GEÔMICA SOBRE TESTES GENÉTICOS.

7. Anand SS, Xie C, Pare G, Montpetit A, Rangarajan S, et al. (2009) Genetic variants associated with myocardial infarction risk factors in over 8000 individuals from five ethnic groups: The INTERHEART Genetics Study. Circ Cardiovasc Genet 2: 16–25.

8. Gladding PA, Legget M, Fatkin D, Larsen P, Doughty R (2020) Polygenic risk scores in coronary artery disease and atrial fibrillation. Heart, Lung and Circulation 29: 634–640.

9. Van Rheenen W, Peyrot WJ, Schork AJ, Lee SH, Wray NR (2019) Genetic correlations of polygenic disease traits: from theory to practice. Nature Reviews Genetics 20: 567–581.

10. Li R, Chen Y, Ritchie MD, Moore JH (2020) Electronic health records and polygenic risk scores for predicting disease risk. Nature Reviews Genetics 21: 493–502.

11. Rodrigues de Moura R, Coelho AVC, de Queiroz Balbino V, Crovella S, Brandão LAC (2015) Meta-analysis of Brazilian genetic admixture and comparison with other Latin America countries. American Journal of Human Biology 27: 674–680.

12. Pena SD, Santos FR, Tarazona-Santos E. Genetic admixture in Brazil; 2020. Wiley Online Library. pp. 928–938.

13. Kehdy FS, Gouveia MH, Machado M, Magalhães WC, Horimoto AR, et al. (2015) Origin and dynamics of admixture in Brazilians and its effect on the pattern of deleterious mutations. Proceedings of the National Academy of Sciences 112: 8696–8701.

14. Carneiro-Proietti AB, Kelly S, Miranda Teixeira C, Sabino EC, Alencar CS, et al. (2018) Clinical and genetic ancestry profile of a large multi-centre sickle cell disease cohort in Brazil. British journal of haematology 182: 895–908.

15. Cubillos-Angulo JM, Arriaga MB, Melo MG, Silva EC, Alvarado-Arnez LE, et al. (2020) Polymorphisms in interferon pathway genes and risk of Mycobacterium tuberculosis infection in contacts of tuberculosis cases in Brazil. International Journal of Infectious Diseases 92: 21–28.

16. Lanas F, Serón P, Lanas A (2013) Coronary heart disease and risk factors in Latin America. Global heart 8: 341–348.

17. Schmidt MI, Duncan BB, e Silva GA, Menezes AM, Monteiro CA, et al. (2011) Chronic non-communicable diseases in Brazil: burden and current challenges. The Lancet 377: 1949-1961.

18. Vrablik M, Dlouha D, Todorovova V, Stefler D, Hubacek JA (2021) Genetics of Cardiovascular Disease: How Far Are We from Personalized CVD Risk Prediction and Management? International Journal of Molecular Sciences 22: 4182.

19. Kim V, Wal Tvd, Nishi MY, Montenegro LR, Carrilho FJ, et al. (2020) Brazilian cohort and genes encoding for drug-metabolizing enzymes and drug transporters. Pharmacogenomics 21: 575–586.

20. Thygesen K, Alpert JS, Jaffe AS, Chaitman BR, Bax JJ, et al. (2019) Fourth universal definition of myocardial infarction (2018). European heart journal 40: 237–269.

21. Sacco RL, Kasner SE, Broderick JP, Caplan LR, Connors J, et al. (2013) An updated definition of stroke for the 21st century: a statement for healthcare professionals from the American Heart Association/American Stroke Association. Stroke 44: 2064–2089.

22. Callum K, Bradbury A (2000) Acute limb ischaemia. Bmj 320: 764–767.

23. Olinic D-M, Stanek A, Tătaru D-A, Homorodean C, Olinic M (2019) Acute limb ischemia: an update on diagnosis and management. Journal of clinical medicine 8: 1215.

24. Harris PA, Taylor R, Minor BL, Elliott V, Fernandez M, et al. (2019) The REDCap consortium: Building an international community of software platform partners. Journal of biomedical informatics 95: 103208.

25. Joseph PG, Pare G, Asma S, Engert JC, Yusuf S, et al. (2016) Impact of a genetic risk score on myocardial infarction risk across different ethnic populations. Canadian Journal of Cardiology 32: 1440–1446.

26. Stram DO (2014) Design, analysis, and interpretation of genome-wide association scans: Springer.

27. Hilbe JM (2016) Practical guide to logistic regression: crc Press.

28. Hosmer Jr DW, Lemeshow S, Sturdivant RX (2013) Applied logistic regression: John Wiley & Sons.

29. Borgan Ø, Breslow N, Chatterjee N, Gail MH, Scott A, et al. (2018) Handbook of statistical methods for case-control studies: CRC Press.

30. Liu Y, Nickleach DC, Zhang C, Switchenko JM, Kowalski J (2018) Carrying out streamlined routine data analyses with reports for observational studies: introduction to a series of generic SAS® macros. F1000Research 7.

31. Bursac Z, Gauss CH, Williams DK, Hosmer DW (2008) Purposeful selection of variables in logistic regression. Source code for biology and medicine 3: 1–8.

32. Royston P, Sauerbrei W (2008) Multivariable model-building: a pragmatic approach to regression anaylsis based on fractional polynomials for modelling continuous variables: John Wiley & Sons.

33. Gönen M (2006) Receiver operating characteristic (ROC) curves. SAS Users Group International (SUGI) 31: 210–231.

34. Walker G, Shostak J (2010) Common statistical methods for clinical research with SAS examples: SAS institute.

35. Lang TA, Secic M (2006) How to report statistics in medicine: annotated guidelines for authors, editors, and reviewers: ACP Press.

36. Romao X, Delgado R, Costa A (2009) An empirical power comparison of univariate goodness-of-fit tests for normality. Journal of Statistical Computation and Simulation.

37. Yap BW, Sim CH (2011) Comparisons of various types of normality tests. Journal of Statistical Computation and Simulation 81: 2141–2155.

38. Yusuf S, Rangarajan S, Teo K, Islam S, Li W, et al. (2014) Cardiovascular risk and events in 17 low-, middle-, and high-income countries. New England Journal of Medicine 371: 818–827.

39. Organization WH (2022) International Classification of Diseases Eleventh Revision (ICD-11). Geneva.

40. Assimes TL, Roberts R (2016) Genetics: implications for prevention and management of coronary artery disease. Journal of the American College of Cardiology 68: 2797–2818.

41. Larsson SC, Burgess S, Michaëlsson K (2017) Association of genetic variants related to serum calcium levels with coronary artery disease and myocardial infarction. Jama 318: 371–380.

42. Wang J, Xu D, Wu X, Zhou C, Wang H, et al. (2011) Polymorphisms of matrix metalloproteinases in myocardial infarction: a meta-analysis. Heart 97: 1542–1546.

43. Roberts R, Chang CC, Hadley T (2021) Genetic risk stratification: a paradigm shift in prevention of coronary artery disease. JACC: Basic to Translational Science 6: 287–304.

44. Lechner K, Kessler T, Schunkert H (2020) Should We Use Genetic Scores in the Determination of Treatment Strategies to Control Dyslipidemias? Current Cardiology Reports 22: 1–9.

45. Weale ME, Riveros-Mckay F, Selzam S, Seth P, Moore R, et al. (2021) Validation of an integrated risk tool, including polygenic risk score, for atherosclerotic cardiovascular disease in multiple ethnicities and ancestries. The American Journal of Cardiology 148: 157–164.

46. Xue Y, Zhang L, Fan Y, Li Q, Jiang Y, et al. (2017) C-reactive protein gene contributes to the genetic susceptibility of hemorrhagic stroke in men: a case-control study in Chinese Han population. Journal of Molecular Neuroscience 62: 395–401.

47. Al Rifai M, Yao J, Guo X, Post WS, Malik S, et al. (2022) Association of polygenic risk scores with incident atherosclerotic cardiovascular disease events among individuals with coronary artery calcium score of zero: The multi-ethnic study of atherosclerosis. Progress in Cardiovascular Diseases.

48. Colares VS, Titan SMdO, Pereira AdC, Malafronte P, Cardena MM, et al. (2014) MYH9 and APOL1 gene polymorphisms and the risk of CKD in patients with lupus nephritis from an admixture population. PLoS One 9: e87716.

49. Castellucci LC, Almeida L, Cherlin S, Fakiola M, Francis RW, et al. (2021) A genome-wide association study identifies SERPINB10, CRLF3, STX7, LAMP3, IFNG-AS1, and KRT80 as risk loci contributing to cutaneous leishmaniasis in Brazil. Clinical Infectious Diseases 72: e515-e525.

